# Prompt identification of the location of gap conduction in the mitral isthmus following vein of Marshall ethanol infusion and endocardial ablation

**DOI:** 10.1101/2024.10.08.24315063

**Authors:** Qiaoyuan Li, Yanguang Li, Zhuo Liang, Tao Zhang, Xu Liu, Dongping Fang, Jin Bai, Jian Li, Fengxiang Zhang, Yunlong Wang

## Abstract

**Background:** Mitral isthmus (MI) gap conduction is common despite ethanol infusion into the vein of Marshall (EI-VOM) and endocardial ablation of the MI. This study aimed to investigate the characteristics of electrograms of the distal coronary sinus (CSd) to guide the identification of the gap location in the MI.

**Methods:** A total of 187 patients who underwent EI-VOM and MI ablation were included in the study. After routine completion of EI-VOM and endocardial MI ablation, the characteristics of the electrogram in the CSd during left atrial appendage (LAA) pacing were analyzed in unblocked MI conduction.

**Results:** Among the 187 patients, 43.3% (81/187) had unblocked MI following EI-VOM and linear lesion creation in the endocardium. In patients with unblocked MI, 84.0% (68/81) showed double potential in the CSd during LAA pacing, among whom 80.9% (55/68) presented with an earlier high-frequency near-field potential (NFP) followed by a low-frequency far-field potential (FFP), suggesting an epicardial gap, whereas 19.1% (13/68) presented with FFP followed by NFP, suggesting an endocardial gap. In patients with single-potential in CSd (16.0%, n = 13), simple activation mapping of the endocardium and CSd revealed the gap location. Intra-CS ablation was necessary in 77.8% (63/81) of the patients, with a mean of 1.3 ± 1.7 sites and 1.1 ± 0.4 min for ablation. Eventually, 95.7 % (179/187) of the patients achieved MI blockage. These findings were confirmed in an external validation cohort, which demonstrated the effectiveness and efficiency of CSd potential-guided gap identification.

**Conclusion:** The characteristics of the electrograms in the CSd could aid in the prompt identification of the gap(s) location of the MI in patients with unblocked MI conduction.

**CLINICAL PERSPECTIVE:** **What is Known:**

Bidirectional mitral isthmus conduction block is challenging despite EI-VOM and endocardial ablation.

**What the Study Adds:**

1. The characteristics of the electrogram at the CSd during left atrial appendage pacing could facilitate prompt identification of gap conduction in mitral isthmus mediated by the epicardium or endocardium.
2. Over 2/3 of patients had an epicardial gap conduction that warranted intra-CS ablation.
3. blockage rate is achievable using the CSd electrogram guided gap identification approach, with efficiency and effectiveness.

**Graphic Abstract:** 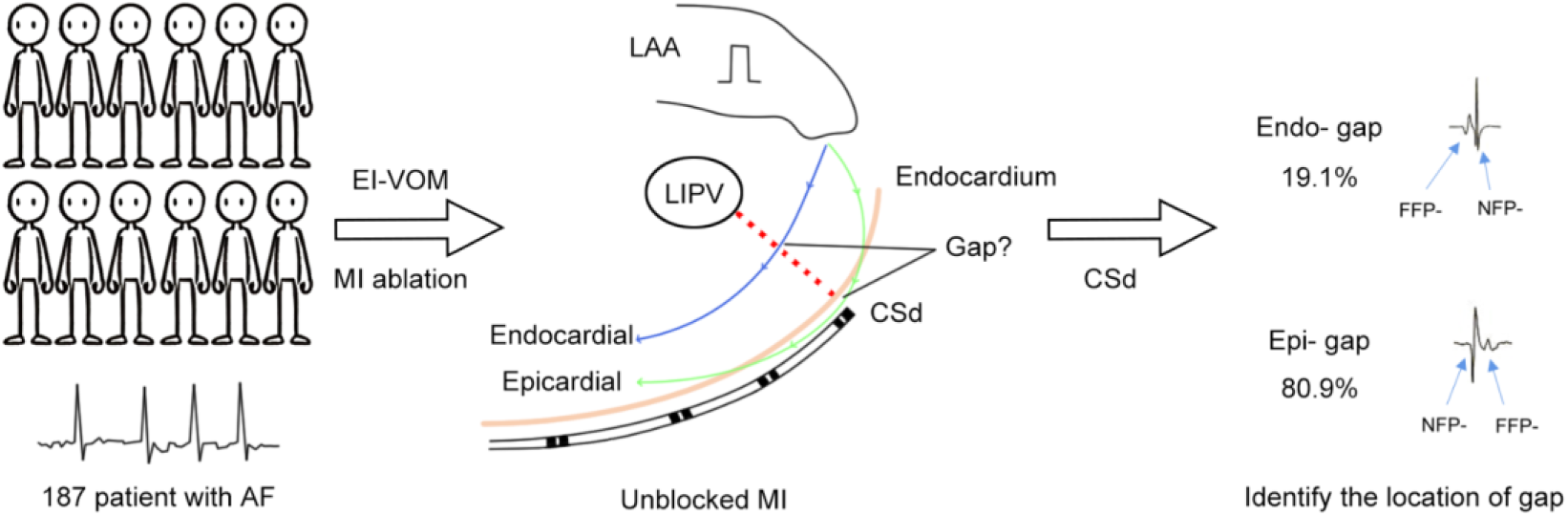

## INTRODUCTION

Mitral isthmus (MI) block is necessary for perimitral atrial tachycardia and atrial compartmentalization beyond pulmonary vein isolation (PVI) in persistent atrial fibrillation. Achieving mitral isthmus (MI) bidirectional conduction block is challenging despite employing ethanol infusion into the vein of Marshall (VOM) (EI-VOM) and endocardial ablation.^[1–5]^ Gap conduction can occur either endocardially or epicardially. Routine endocardial and epicardial ablation with or without systematic mapping are the major approaches employed for managing unblocked MI in clinical practice.^[6]^ Extensive mapping and experimental ablation are ineffective and may increase the risk of tamponade or injury to the circumflex artery.^[7–10]^ Therefore, prompt identification of the location of the gap conduction is necessary for precise ablation.

As a standard landmark, a decapolar mapping catheter is generally placed in the coronary sinus (CS), with the distal electrode located near the MI. The electrogram of the distal CS (CSd) can reflect both epicardial and endocardial activation patterns through near- and far-field potentials (NFP and FFP, respectively). In the present study, we describe a novel approach for the prompt identification of the gap location by analyzing the electrogram characteristics of the CSd during left atrial appendage (LAA) pacing.

## METHODS

### Participants

Consecutive patients who underwent EI-VOM and MI endocardial ablation from Jan 2022 to Dec 2023 were enrolled in this study. The exclusion criteria comprised patients aged < 18 years and those with severe valvular disease, hypertrophic cardiomyopathy, presence of a thrombus in the left atrium, and a history of atrial ablation beyond PVI or tricuspid isthmus block. Written informed consent was obtained from all the patients before the procedure. The study protocol was approved by the institutional review board and adhered to the Declaration of Helsinki guidelines.

### Electrophysiological Study, EI-VOM, and Catheter Ablation

All the participants underwent conscious sedation during the procedure. The procedure was performed using the CARTO-3 mapping system (Biosense Webster, Diamond Bar, CA, USA). A steerable decapolar catheter was placed within the CS with its distal tip located just below the endocardial MI ablation line. Following transseptal access, an intravenous bolus of heparin was administered at 100 U/kg of body weight, targeting an activated clotting time of > 250 s. A multipolar catheter (PentaRay; Biosense Webster, CA, USA) was used for constructing the 3-dimentional structure of the left atrium. The lesion group comprised the EI-VOM, anatomical isthmus linear ablation, and other lesions based on individual patients, including wide antral PVI, roof, and carvotricuspid linear ablation.

For EI-VOM, occlusive CS venography was performed using a wedge balloon infusion catheter through a 6F JR 4.0 guiding sheath (Medtronic) (Figure S1.A). An angioplasty wire (0.014 inch, BMW, Abbot) was advanced into the VOM, followed by a coronary angioplasty balloon catheter (diameter 1.5–2.0 mm and length 8.0–10 mm, depending on the size of the VOM), and advanced in an over-the-wire (OTW) manner into the VOM. With the OTW balloon inflated at 6–10 atm, angioplasty was performed to verify the absence of regurgitation from the VOM and reveal the anatomic characteristics of the VOM (Figure S1.B). Subsequently, 95% ethanol was injected slowly into the VOM. Ethanol was infused starting from the most distal part of the VOM, thereafter, the angioplasty balloon was slightly pulled back proximally, and EI was performed (Figure S1.C–D). This procedure was repeated until the angioplasty balloon was positioned at the VOM ostium. The amount of injected ethanol was 2.0–3.0 mL per infusion with 8–10 mL in total.

Catheter ablation was performed using a 3.5-mm irrigated-tip catheter (ThermoCool SmartTouch SF, Biosense Webster, CA, USA). The power-control mode was set at 40–50 W, with the temperature controlled at 43℃, and normal saline irrigation at 8– 20 mL/min for endocardial ablation. Point-by-point radiofrequency was delivered with an interlesion distance of < 4 mm. The ablation lesion annotation included a catheter stability range of motion of < 2 mm for 4 s and a minimum force of < 8 g for 70% of the time. Each application had a targeted ablation index of 450–600. For intra-CS ablation, the power was set to 20 W, with saline irrigation at 17 mL/min for 20–25 s per ablation site. MI ablation was performed by creating a linear lesion joining the posterior lateral mitral annulus and left inferior pulmonary vein. Ablation catheter stability was optimized using a steerable long sheath (Agilis, St. Jude Medical, St Paul, MN). Upon completion of the MI endocardial linear ablation, electrical or pharmaceutical cardioversion was performed for restoring the sinus rhythm.

### MI Bidirectional Block Verification

The MI bidirectional block was verified using the differential pacing maneuver described by Shah et al.^[11]^ In brief, LAA pacing was performed to observe the CS activation sequence. Clockwise block was defined based on a homogeneous “proximal-to-distal” pattern with CS activation sequence from CS 9–0 to CS 1–2. Differential pacing was conducted in CS 1–2 and CS 3–4 respectively for comparing the activation timing. A counterclockwise block was considered if pacing in CS 3–4 had a shorter stimulation-to-atrial activation timing compared with that of CS 1–2.

### Characteristic Analysis of the CSd Potential in Locating the Gap(s) in the MI Line

In patients with unblocked MI during LAA pacing, the characteristics of the electrograms in the CSd were analyzed and categorized into single and double potentials (Figure 1). Double potential generally exhibits as NFP with high frequency and high amplitude and FFP with low frequency and low amplitude (Figure 1). In cases with an FFP preceding the NFP, the gap was deemed to be located on the endocardial surface, while in those with an NFP preceding the FFP, the gap was deemed to be located on the epicardial surface, i.e., the CS. For patients with a single-potential in the CSd during LAA pacing, the activation timing of the earliest activation site (EAS) in the endocardium just below the MI line was compared with that of the CSd. If the endocardial or CSd potential was observed earlier, the gap was identified as being located in the endocardium or epicardium, respectively. Once the suspected gap was identified, precise ablation was performed at the EAS of either the endocardial or epicardial surface. Figure 1 shows the general flow chart of the study procedure. The method of identifying gap location through the electrocardiogram characteristics of CSd was termed “CSd potential guided gap identification (CSd-GI)” in the present study.

**Figure 1.**
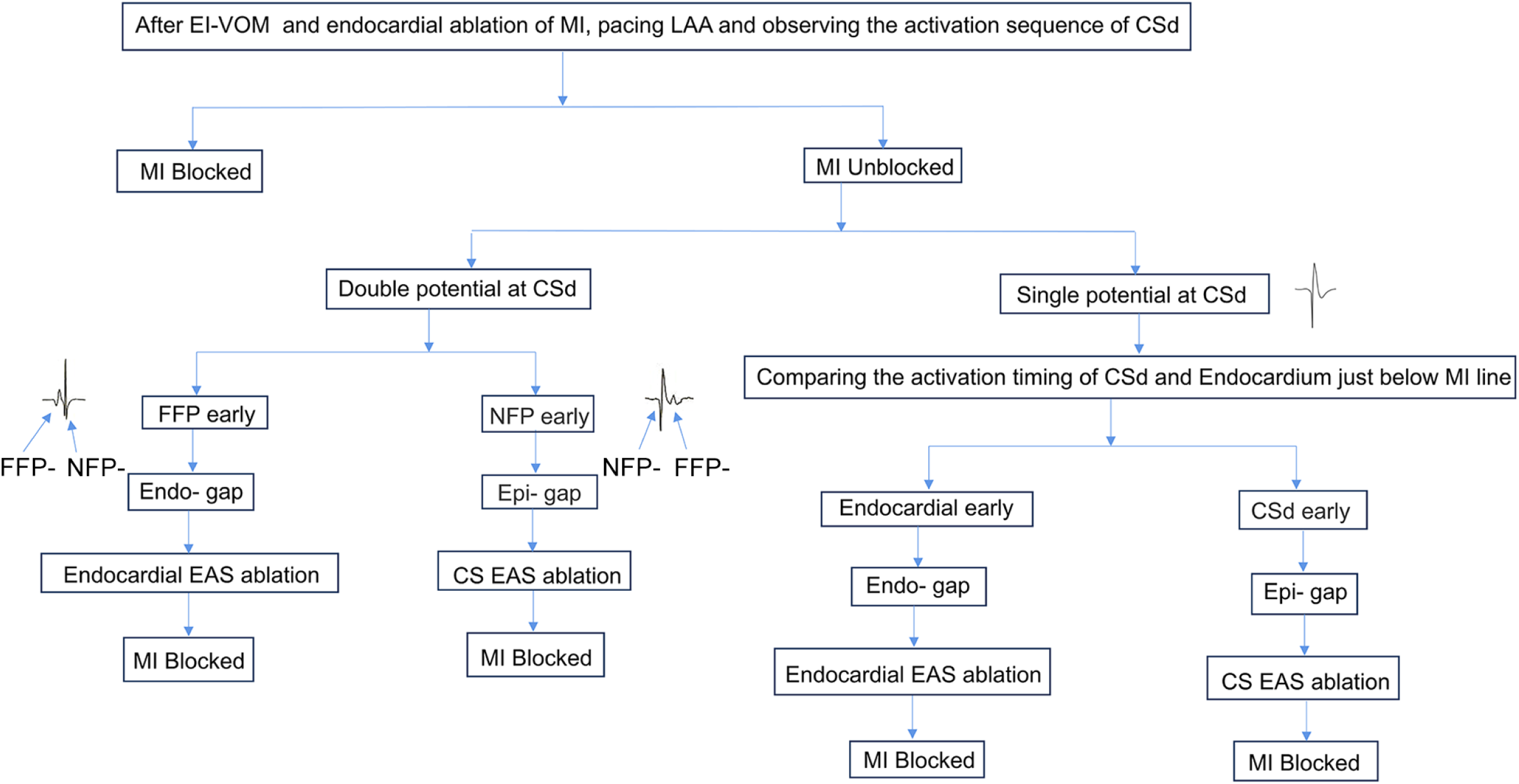
CSd potential guided gap identification in the mitral isthmus line. Abbreviations: CSd, distal coronary sinus; EAS, earliest activation site; EI-VOM, ethanol infusion into the Marshall vein; FFP, far-field potential; LAA, left atrial appendage; MI, mitral isthmus; NFP, near-field potential.

### External Validation

To verify the findings of the present study, external validation was performed in electrophysiological laboratories at three independent centers: Peking University Third Hospital, People’s Hospital of Jiangsu Province, and Chinese People’s Library Army General Hospital. All the patients in the validation cohort underwent EI-VOM and MI ablation. The efficiency and effectiveness of the CSd-GI approach in MI ablation were compared with those of conventional approaches.

### Statistical analysis

Variables are given as mean±SD or percentage, as appropriate. Continuous and categorical data were compared with the Student t test and the χ2 test (or Fisher exact test in case of small sample). *P*<0.05 was considered as significant.

## RESULTS

A total of 187 consecutive patients who underwent MI ablation were enrolled in the present study (mean aged 62.3 ± 8.1 years, 124 [66.3%] male individuals). A total of 167 (89.3%) patients had persistent AF. EI-VOM was successfully performed in 173 (92.5%) patients. In the remaining 14 (7.5%) patients, EI-VOM could not be performed owing to the absence, small size, or tortuosity of the VOM. The patient characteristics are listed in Table 1.

**Table 1.** Patient characteristics (n = 187)

| Items | Value |
| --- | --- |
| Age, years | 62.3 ± 8.1 |
| Male, n (%) | 124 (66.3) |
| Heart rate, bpm | 74 ± 10 |
| Body mass index, kg/m<sup>2</sup> | 27.0 ± 3.5 |
| Brain natriuretic peptide, pg/mL | 223.9 ± 143.9 |
| Estimated glomerular filtration rate, mL/min*1.73 m<sup>2</sup> | 79.7 ± 16.1 |
| Left atrial diameter, mm | 46.8 ± 5.2 |
| Left ventricular ejection fraction, % | 60.7 ± 7.1 |
| Duration of atrial fibrillation, months* | 31.4 ± 24.2 |
| Time from diagnosis to procedure, months | 43.5 ± 29.3 |
| Hypertension, n (%) | 105 (56.1) |
| Diabetes mellitus, n (%) | 33 (17.6) |
| Coronary artery disease, n (%) | 31 (16.6) |
| Peripheral vascular disease, n (%) | 19 (10.2) |
| Cerebrovascular disease, n (%) | 10 (5.3) |
| Obstructive sleep apnea syndrome, n (%) | 16 (8.6) |
| CHA<sub>2</sub>DS<sub>2</sub>-VASc score | 2.8 ± 1.4 |

\* Patients with a history of atrial fibrillation (n = 167)
CHA<sub>2</sub>DS<sub>2</sub>-VASc score = congestive heart failure, hypertension, age, diabetes mellitus, prior stroke or TIA or thromboembolism, vascular disease, age, sex category

After EI-VOM and endocardial MI linear ablation, 106 (56.7%) patients achieved MI blockage. Of the remaining 81 patients with unblocked MI, 68 (83.9%) showed double potential in the CSd during LAA pacing. Among those with double potentials, 80.9% (55/68) showed earlier NFP followed by FFP (Figure 2.a1–a2), while 19.1% (13/68) presented with earlier FFP followed by NFP (Figure 2.b1–b2). By comparing the activation timing of the NFP in the CSd and endocardial surface just below the MI line, we found that the gap was located at the endocardium of the MI for those with FFP proceeding NFP in the CSd. For those with NFP proceeding FFP in the CSd, we found that the gap was located within the epicardium. Ablation of the gap abolished MI conduction (Figure 2.a3–a4 and b3–b4).

**Figure 2.**
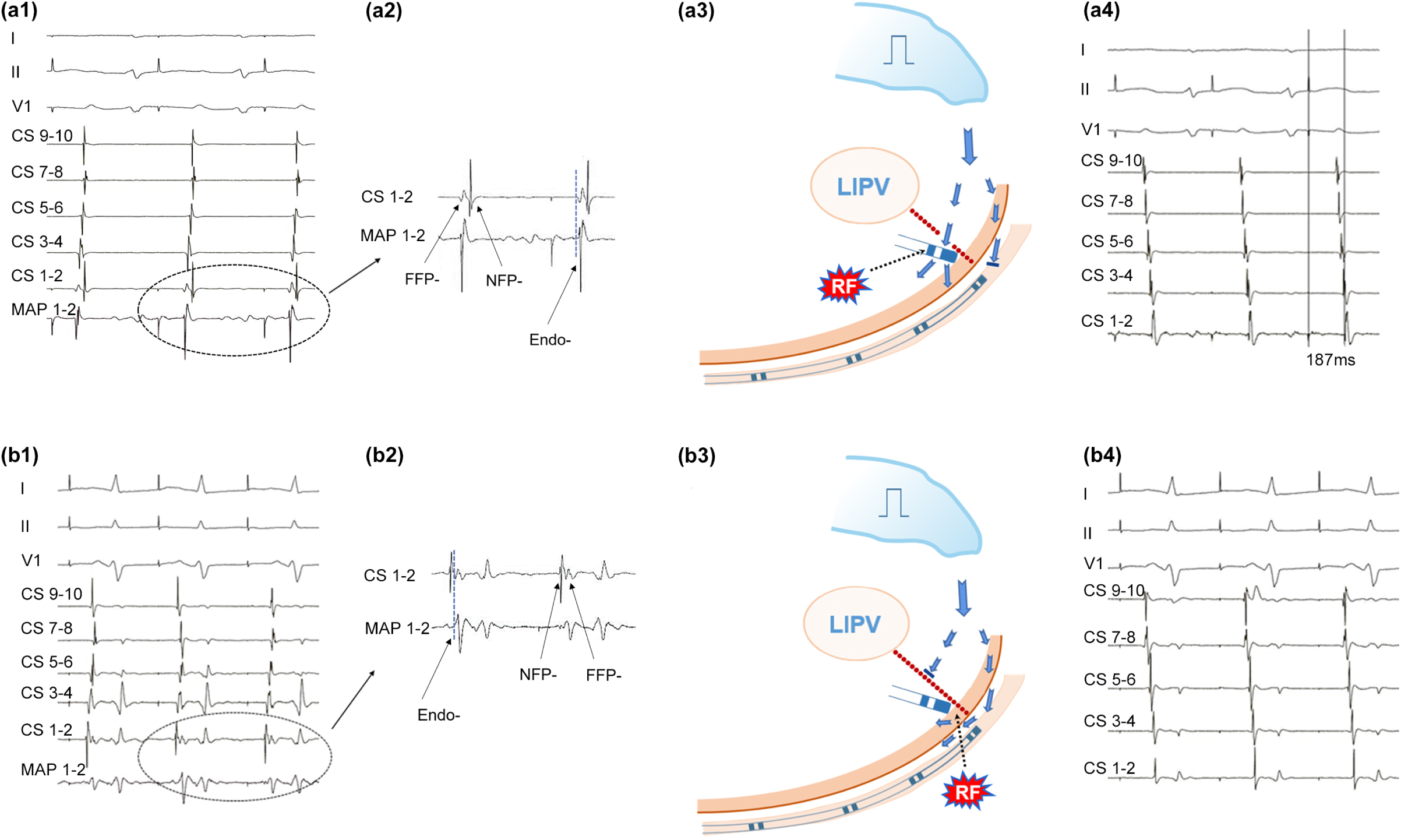
Characteristics of the electrogram in the CSd revealing the gap location of the mitral isthmus. a1: During left atrial appendage (LAA) pacing, a double potential was recorded at the distal coronary sinus (CSd) (CS 1–2), i.e., a low-frequency far-field potential (FFP) followed by a high-frequency near-field potential (NFP). a2: Endocardial mapping just below the mitral isthmus (MI) indicated simultaneous activation between the FFP of the CSd and earliest activation site (EAS) at the endocardium, suggesting that the gap was located in the endocardial surface of the MI. a3: Schematic illustrating this phenomenon; ablation was performed at the EAS of the endocardium (RF). a4: Endocardial ablation of the EAS abolished MI conduction. b1: During LAA pacing, a double potential was recorded at the CSd, i.e., a high-frequency NFP followed by a low-frequency FFP. b2: Comparison between the CSd and endocardial MI indicated that the gap was located on the epicardium of the MI, i.e. the CS. b3: Schematic diagram illustrating this phenomenon; intra-CS ablation was performed (RF). b4: CSd ablation abolished MI conduction. Abbreviations: LIPV=left inferior pulmonary vein; RF=radio frequency ablation.

In the remaining 16.1% (n = 13) of the patients with a single potential at the CSd during LAA pacing, comparison of the activation timing of the endocardium just below the MI line and that of the CSd (Figure 3. b1) revealed the gap location in the endocardium in 5 (38.5%) patients (example: site 1 in Figure 3.b). Ablation of the endocardium in the EAS successfully abolished MI conduction (Figure 3.b4). In the remaining 8 (61.5%) patients, the EAS was located in the epicardium. Ablation of the EAS in the epicardium abolished MI conduction (Figure 3.a4). The mean number of ablated sites (duration) in the endocardium and epicardium for MI block was 2.6 ± 1.4 (1.1 ± 0.8 min) and 1.4 ± 0.6 (0.4 ± 0.3 min), respectively.

**Figure 3.**
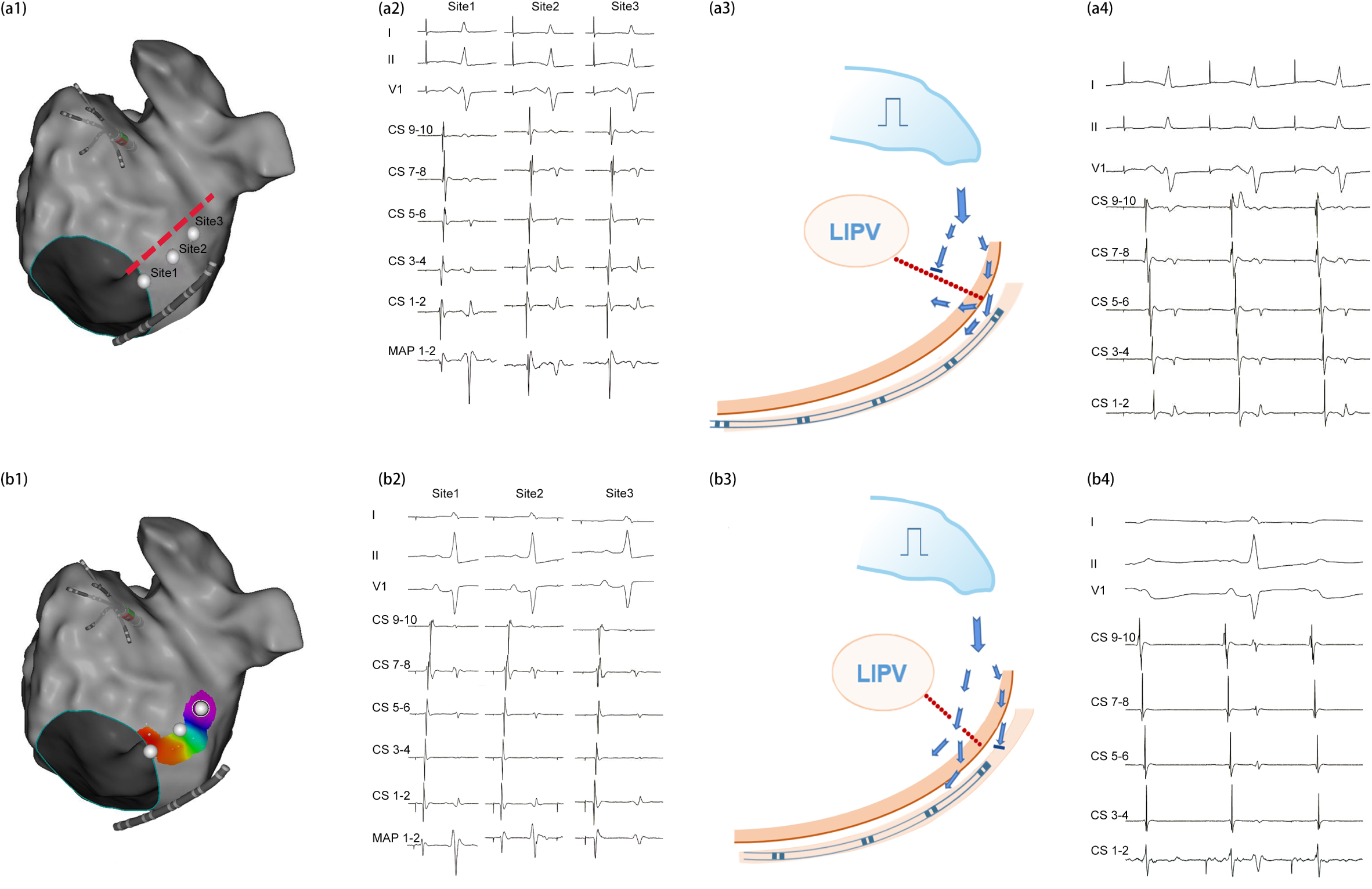
Activation mapping comparing the sequence of the endocardium and CSd. a1: Simple activation mapping of the endocardium and comparison of the endocardial earliest activation site (EAS) with that of the distal coronary sinus (CSd). a2: Comparison of the activation sites indicates earlier activation in the CSd than in the endocardial EAS, suggesting an epicardial gap. a3: A schematic diagram illustrating this phenomenon; intra-CS ablation was conducted. a4: Intra-CS ablation abolished MI conduction. b1: Activation mapping suggests earlier activation of all the endocardial activation sites than the CSd, suggesting an endocardial gap. b2: The EAS is located at site 1. b3: A schematic diagram illustrating this phenomenon. Ablation was conducted in the endocardium EAS. b4: Ablation of the endocardial EAS (site 1) abolished MI conduction.

Electrocardiogram changes were observed in 12.3% (10/81) of the patients during endocardial or epicardial ablation, indicating the coexistence of both endocardial and epicardial gaps (Figure 4). In an unblocked MI (Figure 4), ablation in the endocardium resulted in slowed local conduction, with the FFP being delayed, and the formation of a double potential at the CSd, with the NFP presenting earlier than the FFP (Figure 4.a3). Further ablation of the epicardium resulted in complete MI blockage (Figure 4.a4). Pseudo-blockage occurred after the completion of epicardial ablation in 7.4% (6/81) of the patients (Figure 4.b3). During LAA pacing, the activation sequence of the decapolar catheter in the CS was CS 9–0 to CS 1–2 as reflected by the NFP. Fine analysis of the electrograms in the CS indicated the presence of a double potential in the distal section of the decapolar catheter, with a clockwise activation sequence from CS 1–2 to CS 5–6 as reflected by the FFP, suggesting endocardial conduction through the MI line (Figure 4.b3). Further ablation of the endocardium abolished MI conduction (Figure 4.b4).

**Figure 4.**
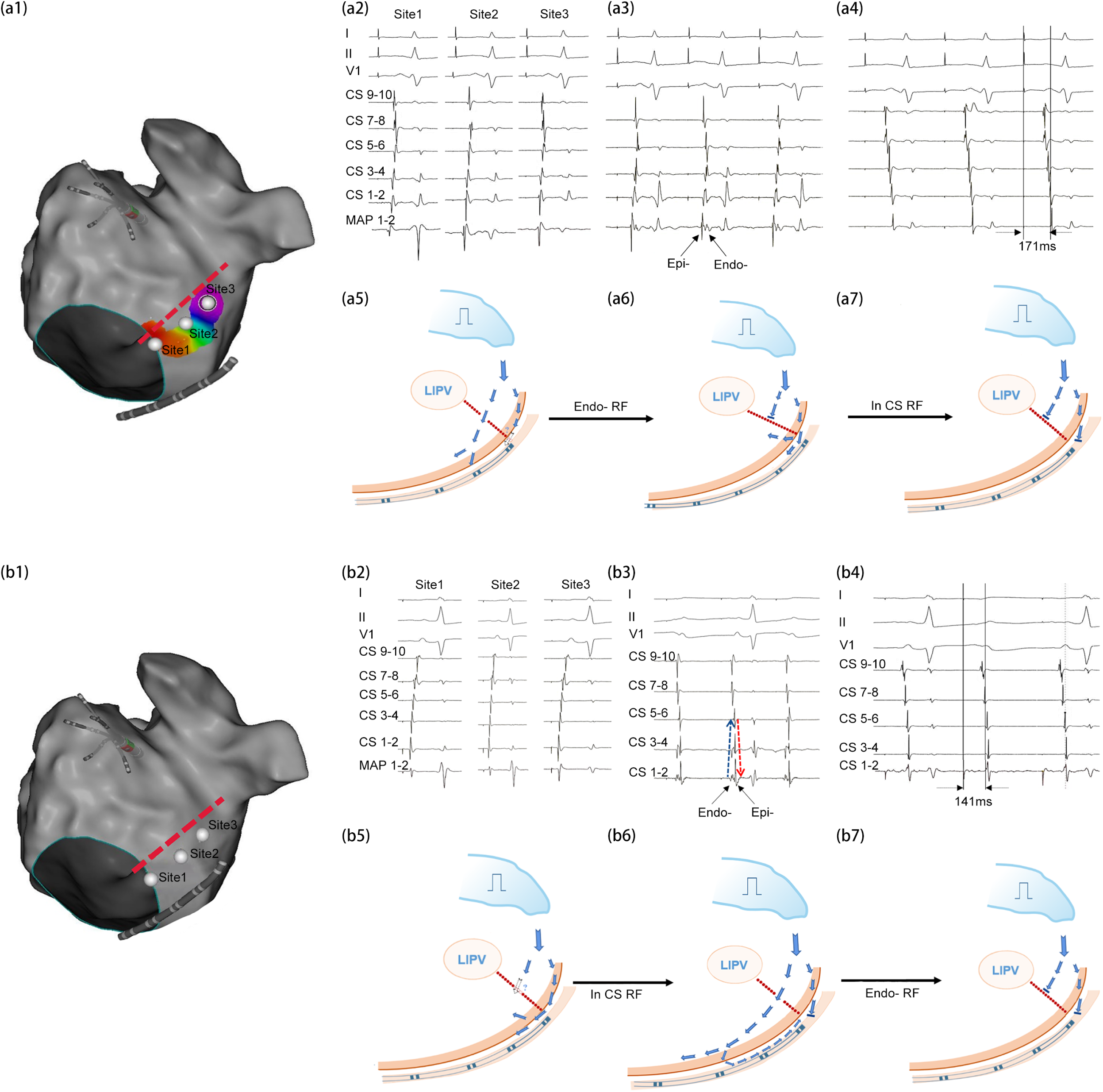
CSd potential changes during ablation indicated coexistence of both endocardial and epicardial gaps. a1–a2: Activation mapping to determine the earliest activation site (EAS) on the endocardial surface and compare the endocardial EAS with the activation timing of the distal coronary sinus (CSd). a3: Ablation of the endocardium resulted in the occurrence of a far-field potential (FFP) attached to the near-field potential (NFP) at the CSd. a4: Further ablation of the epicardium resulted in complete mitral isthmus (MI) blockage. a5: A schematic diagram illustrating a2 phenomenon. a6: A schematic diagram illustrating a3 phenomenon. a7: A schematic diagram illustrating a4 phenomenon. b1–b2: Activation mapping indicated that activation in the CSd occurred earlier than that in the endocardium. b3: Ablation of the epicardium resulted in a delay in the NFP and presence of an FFP at the CSd. b4: Further ablation of the endocardium resulted in complete MI block. b5: A schematic diagram illustrating b2 phenomenon. b6: A schematic diagram illustrating b3 phenomenon. b7: A schematic diagram illustrating b4 phenomenon.

Characteristic electrocardiogram findings in the CSd were also noted in guiding gap identification (Figure 5) in perimitral atrial tachycardia. If the NFP occurred earlier than the FFP, the gap was located at the epicardium, whereas if the FFP preceded the NFP, the gap was located at the endocardium.

**Figure 5.**
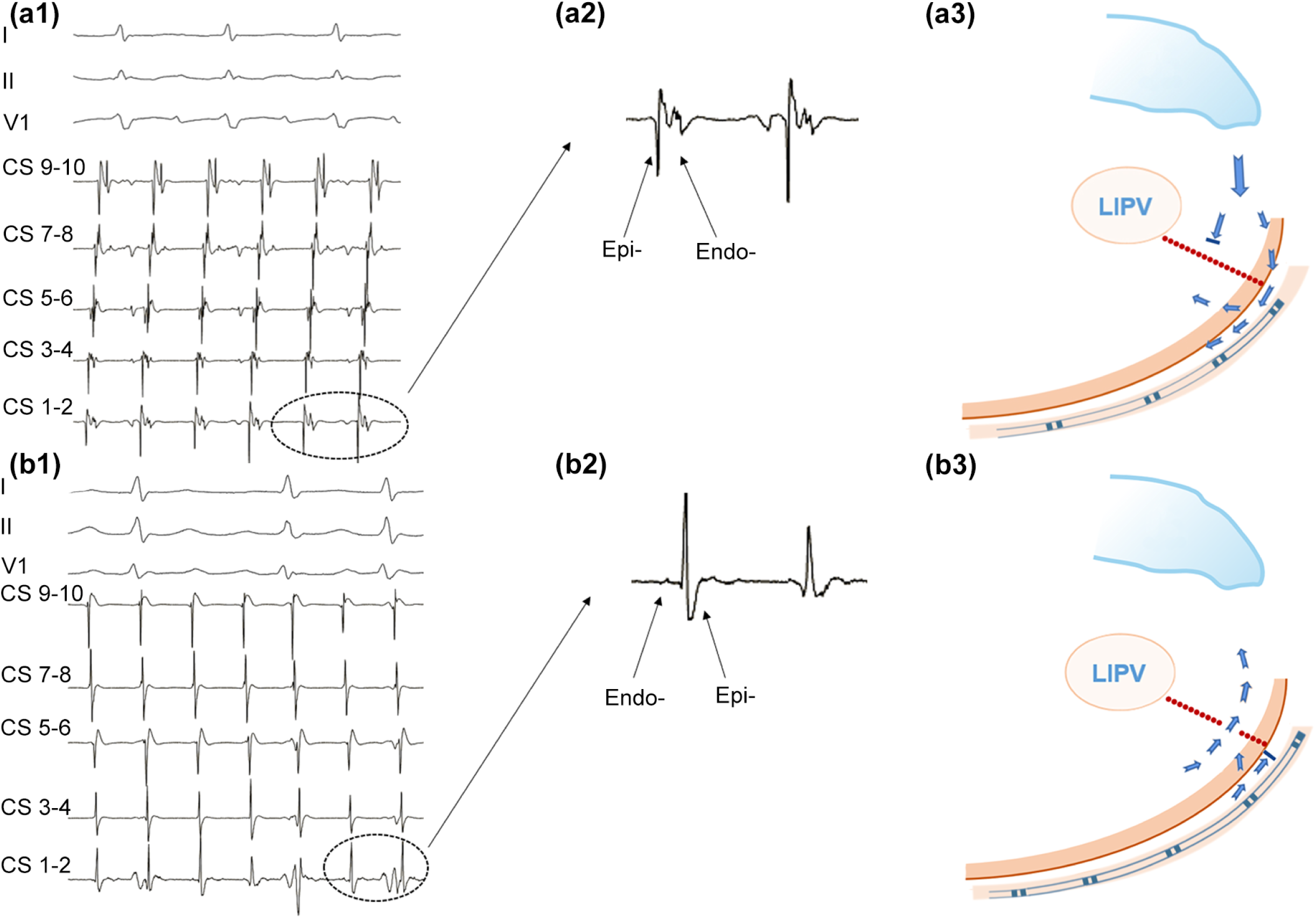
CSd potential characteristics for guiding gap identification in perimitral atrial tachycardia. a1: Clockwise perimitral atrial flutter. a2: Double potential was observed in the distal coronary sinus (CSd) with a near-field potential (NFP) preceding the far-field potential (FFP), suggesting that gap conduction was located at the epicardium. a3: Schematic illustration of this phenomenon. b1: Counterclockwise perimitral atrial flutter. b2: Double potential was present in the CSd with the FFP preceding the NFP, suggesting that gap conduction was located at the endocardium. b3: Schematic illustration of the phenomenon.

With the use of the CSd-GI approach, 95.7% (179/187) of the patients achieved MI blockage in the present study. The total ablation time in the endocardium of the MI was 4.3 ± 3.6 minutes. After completion of the endocardial MI line, intra-CS ablation was necessary in 77.8% (63/81) of the patients, with a mean of 1.3 ± 1.7 sites and 1.1 ± 0.4 minutes for ablation. The mean number of ablated sites (duration for gap conduction elimination) in the endocardium and epicardium were 3.4 ± 1.6 (1.3 ± 0.7 min) and 1.7 ± 0.9 (0.6 ± 0.4 min), respectively.

In patients with an epicardial gap (n = 63), 79.4 (50/63) of the successfully ablated sites were located near the ostium of the VOM, 12.7% (8/63) were located in the proximal section of the CS, and 7.9% (5/63) were located in the great cardiac vein (GCV) (Figure 6). Among the epicardial gaps, 32.2% of the successfully blocked sites were located in the free wall of the CS-GCV muscular structure with a contact force vector toward the free wall.

**Figure 6.**
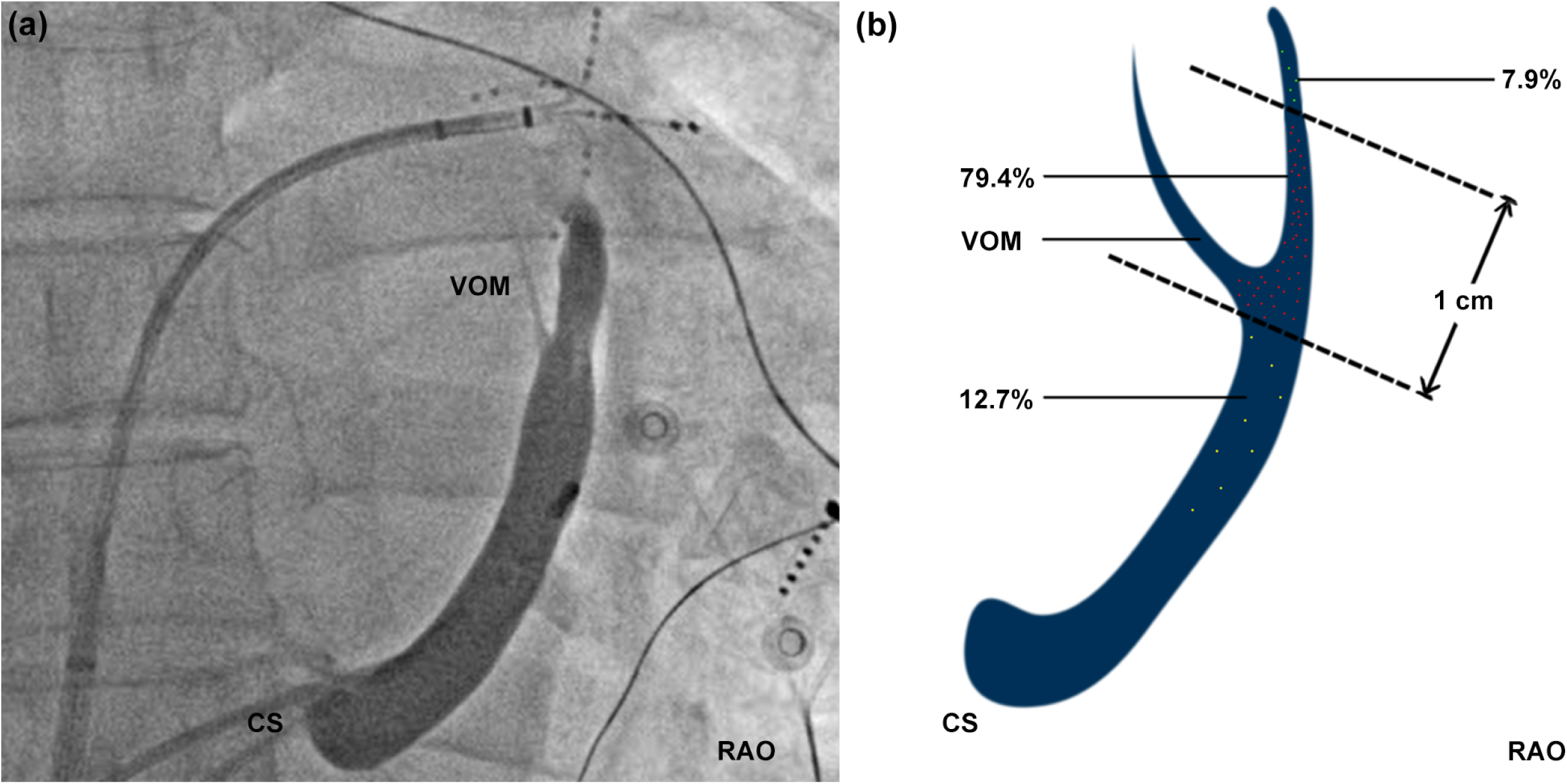
Relationship between the ablation target within the coronary sinus and great cardiac vein. a: The Marshall vessel under contrast imaging. b: 79.4% were located within 1cm of the Marshall vessel. 7.9% located more than 1cm away from the distal end of Marshall vessel, while 12.7% located near the Marshall vessel.

### External Validation

Whether more efficient, effective, and successful MI blockage could be achieved using the CSd-GI approach was validated in an external cohort. A total of 31 patients with unblocked MI treated using the CSd-GI method (CSd-GI group) were enrolled at three external electrophysiological centers. For comparison, 42 patients who underwent conventional MI ablation approaches were retrospectively analyzed using relevant data. All the patients underwent EI-VOM before MI ablation. The baseline characteristics of the two patient groups are shown in the supplementary file (Table S1). The CSd-GI group demonstrated a higher MI blockage rate, fewer ablation sites, and a shorter ablation duration in both the endocardium and epicardium (P < 0.05).

## DISCUSSION

The present study investigated the role of CSd electrograms in guiding gap identification in the MI. The main findings are as follows: 1) After initial completion of EI-VOM and endocardial ablation of the MI, only half the cases achieved MI blockage; 2) the characteristics of the electrogram at the CSd could facilitate prompt identification of gap conduction mediated by the epicardium or endocardium; 3) over 2/3 of the patients had an epicardial gap conduction that warranted intra-CS ablation; 4) through the CSd-GI approach, a high MI blockage rate was achieved (95.7%); and 5) the CSd-GI approach was validated in an external cohort, demonstrating improved efficiency and effectiveness with fewer ablation sites and shorter durations when compared with those using conventional approaches.

MI ablation is necessary for perimitral atrial tachycardia and atrial compartmentalization beyond PVI; however, the blockage rate was unsatisfactory. Although this rate has increased significantly with the use of EI-VOM, achieving MI blockage is challenging in cases warranting extensive activation mapping and ablation. In the study by Pambrun et al., the initial blockage rate was 51% following EI-VOM and endocardial ablation.^[12]^ Thus, additional gap identification is warranted for achieving higher blockage rates.^[12, 13]^. In the present study, we found that after initial completion of EI-VOM and endocardial ablation, only 43.3% of patients achieved MI blockage, which could be attributed to insufficient lesion in VOM and extensive CS-GCV connections.

Conventional approaches include routine intra-CS ablation and repeated ablations of the endocardium, which result in unnecessary lesions and risk of complications. Kawaguchi et al. reported the need for epicardial ablation through the CS-GCV in nearly half of the cases for eliminating trans-epicardial MI conduction.^[12, 14]^ However, distinct indications for epicardial ablation have not been elucidated. Identification of the gap location, either endocardially or epicardially, in unblocked MI is crucial for targeting precise ablation sites and avoiding unnecessary lesions, especially in the CS. Alternatively, thorough endocardial and epicardial high-density activation mapping can identify the location of gap conduction, which is generally a laborious maneuver, can identify the location of gap conduction.^[12]^ In the present study, we found that the characteristics of the electrogram in the CSd could facilitate prompt identification of the gap location.

Electrograms inside the CS generally consist of two components: far-field left atrial potential and near-field CS musculature potential.^[15]^ In the present study, with the distal decapolar catheter in the CS placed just beneath the endocardial MI line, both the endocardial and epicardial activation sequences could be recorded during LAA pacing. We found that a double potential at the CSd was common, with the far-field component representing left atrial activation (endocardial) and the near-field component representing CS musculature activation (epicardial). The FFP and NFP sequences revealed whether the advanced activation site was located in the endocardium or epicardium, respectively. In those with a double potential at the CSd, if the NFP preceded the FFP, gap conduction was located in the epicardium, whereas if the FFP preceded the NFP, gap conduction was located in the endocardium. Therefore, by analyzing the characteristics of the local electrogram near the MI, i.e., CSd potential, a laborious activation mapping could be waived.

Occasionally, only single-potential could be recorded at the CSd, probably due to the simultaneous activation of both the endocardium and epicardium, with a fusion of the NFP and FFP, or because the distal CSd was adjacent to the free wall of the CS-GCV away from the endocardium, resulting in loss of endocardial activation recording. For those with only a single potential at the CSd, a simple comparison of the EAS in the endocardium just beneath the MI line and CSd can identify the gap location. In some cases, the manipulation of the decapolar catheter with displacement of its tip could exhibit a double potential. Interestingly, we found that for those with a single potential at the CSd, ablation in the endocardium or epicardium can result in splitting of this single potential and formation of a double potential, which has been reported in a previous study investigating the role of EI-VOM in MI conduction.^[14]^ Furthermore, endo-ablation can lead to a delay in the FFP, whereas epi-ablation can lead to a delay in the NFP. This phenomenon further confirms our theory of the significance of double potential in guiding gap identification in MI.

Using the CSd-GI approach, the MI blockage rate was 95.7%. Furthermore, in the external validation cohort comprising the conventional MI ablation and CSd-GI approaches, a higher MI blockage rate, fewer ablation sites, and a shorter ablation duration in both the endocardium and epicardium were observed in the CSd-GI group. Prompt identification of the gap location can eliminate unnecessary mapping and ablation, especially intra-CS ablation, which is associated with underlying injury to the circumflex artery and CS stenosis.^[8]^ For patients with unblocked MI, EI-VOM was not successfully performed, which might emphasize the significance of EI-VOM in eliminating complex trans-MI conduction.

Regarding the distribution of successful blockage sites in the epicardium, we found that the majority of the gap locations were located proximal or distal to the ostium of the VOM. Moreover, more proximal (middle-CS) and distal (middle-GCV) gaps also exist, which vary from those reported in previous studies.^[12]^ This difference may be attributed to the variations in the relationship between the ostium of the VOM and MI lines. If the ostium of the VOM is near the proximal CS and away from the MI, then the gaps might be located in the middle GCV, and vice versa. Therefore, anatomical variations should be considered when searching for the location of gap conduction.

### Limitations

The present study had some limitations. First, the CSd-GI approach cannot be employed in patients with insufficient lesions in the EI-VOM and abundant trans-epicardial connections, which are relatively scarce. Second, a preliminary exploration of the connection between the left atrium and surrounding fibers of the CS-GCV was not performed as it was not the major focus of the present study. Third, our sample size was relatively small. Fourth, long-term follow-up data for observing maintenance of a durable MI conduction block are lacking.

## Conclusion

Gap identification was still required in more than half of the patients following EI-VOM and initial endocardial ablation. The CSd-GI approach can help in the prompt identification of the location of the gap conduction, thereby serving as an efficient and effective method for improving the MI blockage rate.

## Data Availability

All the date were availability.

## NON-STANDARD ABBREVIATIONS AND ACRONYMS

CS: coronary sinus
CSd: distal coronary sinus
CSd-GI: CSd electrogram guided gap identification
EAS: earliest activation site
EI: ethanol infusion
FFP: far-field potential
GCV: great cardiac vein
LAA: left atrial appendage
LIPV: left inferior pulmonary vein
MI: mitral isthmus
NFP: near-field potential
OTW: over-the-wire
PVI: pulmonary vein isolation
RF: radiofrequency ablation
VOM: vein of Marshall

## ACKNOWLEDGMENTS

Drs Wang YL accessed and verified the underlying data in this study and take responsibility for the accuracy of the data analysis. Drs Li QY, and Li YG wrote this article. Drs Liang, Zhang, Fang DP, and Liu contributed to the article revision. Drs Bai, Li, and Zhang FX performed the external validation. Drs Wang YL designed the study and interpreted results. All authors read and approved the final article.

## SUPPLEMENTAL MATERIAL

Figure S Ethanol infusion in to the vein of Marshall and correlated voltage changes thereafter

a: Application of 6F JR4.0 Guidance in coronary sinus proximal angiography, clear display of vein of Marshall (VOM) manifestation. b: Guide the wire to the distal of VOM, and insert a balloon along it to the middle section of the VOM. After inflation of the balloon, re-imaging confirms that VOM has been completely sealed by the balloon and there is no reflux. c: Slowly insert 8-10ml of 95% alcohol into VOM (EI-VOM). d: Re-contrast, confirming the presence of contrast agent retention along VOM path. e1-e2: Comparison of left atrial substrate changes before and after EI-VOM in a posterior anterior position. f1-f2: Left side comparison of changes in left atrial substrate before and after EI-VOM.

Table S1 Characteristics of external validation cohort

